## Supplementary material for "COVID-19 vaccine for people who live and work in prisons worldwide: A scoping review": Search terms

**Appendix 1**

The following subject headings were included as exploded terms for *prisons/prisoners* if present in the database: prison (EMBASE), prisons (PsycEXTRA, Medline, PsycINFO), prisoner (EMBASE), prisoners (PsycEXTRA, Medline, Global Health, CINAHL, psycINFO), prisoners of war (PsycEXTRA), prisoner abuse (PsycEXTRA, PsycINFO), criminal offenders (PsycEXTRA), Criminal Rehabilitation (PsycEXTRA), correctional institutions (Global Health), offender (EMBASE), prisoner nursing (EMBASE), correctional facilities personnel (CINAHL), correctional health services (CINAHL), correctional facilities (CINAHL), correctional health nursing (CINAHL), prison personnel (PsycINFO). This search was performed in all databases looking only at the title and abstract of articles.

These terms were combined with a text word search for the following: prison* OR inmate OR inmates OR staff OR officers OR personnel OR workforce OR workers OR jail OR gaol* OR correction* facilit* OR penitentiar* OR penal institut* OR detention camp* OR custod* OR concentration camp* OR incarcerate* OR imprison* OR correctional setting* OR detain* OR detention* OR correction* cent* OR compulsory drug detention OR compulsory drug treatment OR compulsory rehabil* OR "re‐education through labo*" OR "long‐term detention" OR labo* camp*. This search was performed in all databases looking only at the title and abstract of articles.

The following subject headings were included as exploded terms for *COVID-19* if present in the database: COVID-19 (CINAHL). These terms were combined with a text word search for the following: coronavirus* OR coronovirus* OR Wuhan OR "2019-nCoV" OR COVID OR “COVID-19” OR “CORVID-19” OR “CONVID-19” OR "WN-CoV” OR “HCoV-19” OR CoV OR "2019 novel" OR ncov OR "SARS-CoV-2" OR SARSCov19 OR ncov*wuhan OR “novel betacov” OR “novel betacoronavirus”. This search was performed in all databases looking only at the title and abstract of articles.

The following subject headings were included as exploded terms for *vaccination* if present in the database: vaccine (EMBASE), vaccination (EMBASE, MEDLINE), immunisation (EMBASE). These terms were combined with a text word search for the following: Vaccin* OR immun*
