## Supplementary material for "COVID-19 vaccine for people who live and work in prisons worldwide: A scoping review": Abbreviated data charting analysis

**Appendix 2 – Abbreviated data charting form**

| Author, month and year, title | Study design | Population described or studied | Key findings and recommendations related to COVID-19 vaccination in prisons |
| --- | --- | --- | --- |
| Adebisi et al., April 2021, “Vaccinate people in Africa’s prisons against COVID-19” | Correspondence | African prisons | The authors argue that Africa’s prison population should be a priority group to receive COVID-19 vaccines because of overcrowding, poor ventilation in cells, and unsanitary conditions. Prisons have high incidence of chronic diseases such as hypertension and diabetes. These increase the likelihood of poor outcomes from COVID-19 infection, especially as sufferers in prison often have limited access to healthcare. Prisons can also drive transmission via staff and visitors. |
| Amnesty International, March 2021, “Forgotten behind bars: Covid-19 and prisons” | Report | Worldwide | Amnesty International state that the lack of clarity about vaccination plans, policies, and treatment of incarcerated populations should be a global concern. While some countries have already adopted policies that prioritise prison populations and staff to receive vaccines, many others, including high-income countries, remain silent or ambiguous about their plans. The report urges countries to draft national vaccination plans that prioritise people who live and work in prisons. This group is stated to be at particular risk as their confined conditions do not allow them to physically distance. |
| Barnert et al., June 2021, “Ten Urgent Priorities Based on Lessons Learned from More Than a Half Million Known COVID-19 Cases in US Prisons” | Research and analysis | US prisons | With the rise in availability of vaccines, health experts have called for prison residents and staff to receive priority vaccination. However, viral mutations, lags in vaccine distribution, and vaccine hesitancy mean that COVID-19 mitigation techniques, such as limiting transfers between facilities and improving prison ventilation systems, will be required for the foreseeable future. |
| Barsky et al., April 2021, “Vaccination plus Decarceration — Stopping Covid-19 in Jails and Prisons” | Perspective | US prisons | The authors provide examples from various US jurisdictions of how prioritisation of people in prisons in state vaccination plans has been overturned by state politicians, who disregarded both epidemiological reality and the likelihood of downstream harm to the general public.  The authors also anticipate high rates of vaccine hesitancy among staff and especially among incarcerated people, who have been offered little to no educational material regarding COVID-19 vaccines and have abundant reasons to distrust any official recommendations, given long-standing violations of basic human rights and histories of abuse in US carceral facilities. |
| Berk et al., April 2021, “Why we vaccinate incarcerated people first” | Editorial | US prisons | The authors argue that vaccine allocation should follow the epidemiology and be first deployed in settings where vaccination would have the biggest impact, i.e. in places that have seen high case numbers and high disease severity. Jails and prisons meet these criteria. Vaccinations within prisons would reduce healthcare costs and allow access to a targeted population housed in one place. The right to receive vaccination is also a constitutional right in the US. |
| Biehl et al., June 2021, “Supreme Court v. Necropolitics: The Chaotic Judicialization of COVID-19 in Brazil” | Special Section | Brazilian population, including people in prisons | This paper highlights the biopolitical role of the Brazilian judiciary during the pandemic, probing the effect that COVID-19 judicialisation had on the separation of powers and on human rights and accountability. The work examines how the Brazilian Supreme Court advanced rights protection and implementation of evidence-based interventions to ensure that people in prisons benefit from vaccination programmes. |
| Blumberg et al., July 2021, “Mitigating outbreaks in congregate settings by decreasing the size of the susceptible population” | Empirical | 35 California state prisons | Interventions that pre-emptively reduce the susceptible population have a greater than linear effect on outcomes. Under the model assumptions made, reducing the size of the susceptible population by 20% reduced overall disease burden by 47%. Measures that would decrease the susceptible population include depopulation (for example through decarceration), and vaccination. |
| Brathwaite et al., June 2021, “High COVID-19 death rates in prisons in England and Wales,  and the need for early vaccination” | Spotlight | Prisons in England and Wales | In the UK, people in prisons are currently being offered COVID-19 vaccines in line with general national prioritisation criteria, i.e. based on age and presence of long-term conditions. The authors argue that it can be difficult to identify eligible people in prisons due to poor clinical coding and limited interaction with health services, both prior to and during time in prison. Many people in prisons are therefore considered low priority by default. The paper also reports on vaccine hesitancy in prisons among younger people and those from ethnic minorities. It concludes that simultaneous vaccination of prison populations, including staff, could reduce mistrust and expedite a return to normal functioning. |
| Brinkley-Rubinstein et al., July 2021, “Breakthrough SARS-CoV-2 Infections in Prison after Vaccination” | Correspondence | US prisons | The authors question why COVID-19 vaccine rollout has varied across prison and jail populations despite the Centers for Disease Control and Prevention, the National Academy of Medicine, and the American Medical Association all recommending prioritisation. The authors also note that few studies have been conducted on the effectiveness of vaccination efforts in congregate housing. Of those that have been conducted, most have been in skilled nursing facilities, where vaccine effectiveness has been reported to be 63 to 64%. |
| Cervin, March 2021, “Justice required: Vaccination in Canadian prisons” | President’s Message/Editorial | Canadian prisons | More than half of those detained in Canada are on remand awaiting trial. These individuals have not been convicted of a crime and might return to the community once their hearing has been held. Of those sentenced, median incarceration time is just 30 days. Prisons are congregate settings where people with high rates of comorbidity reside before returning to the general population. The author argues that if the Canadian government is serious about treating its citizens equitably and upholding their human rights, it must ensure swift access to vaccinations for people in prisons. |
| Chapman et al., June 2021, “Comparison of COVID-19 vaccine prioritization strategies” | Original research | Demographic and location data  on 28,175 COVID-19 deaths in California up to 3-30^th^ December 2020 across all locations including prisons. | The study highlights the strength of age-based targeting of vaccination as a strategy for averting COVID-19 deaths and the potential benefit of targeting by other risk factors. The authors argue that because many countries still have low vaccination coverage, ensuring that vaccines reach those most at risk of poor health outcomes should remain a focus of the vaccine rollout. |
| Chin et al., June 2021a, “Covid-19 Vaccine Acceptance in California State Prisons” | Correspondence | California state prisons | Younger and healthier prison residents were found to be less likely to accept vaccines than older and medically vulnerable residents. Acceptance was markedly lower among Black residents, a finding that may reflect increased mistrust in correctional authorities and clinicians among this group, or a lack of access to reliable information on vaccine safety and efficacy. However, the study also showed that a substantial proportion of residents who initially declined a first dose later accepted a re-offer, indicating that perceptions towards vaccines could change. |
| Chin et al., August 2021b, “Effectiveness of COVID-19 Vaccines among Incarcerated People in California State Prisons: A Retrospective Cohort Study” | Original research | California prisons | Consistent with results from randomised trials and observational studies in other populations, mRNA vaccines were highly effective in preventing SARS-CoV-2 infections among incarcerated people. The authors argue that prioritising incarcerated people for vaccination, redoubling efforts to boost the rates of initial vaccination, and continuing other ongoing mitigation practices are essential for preventing COVID-19 in this disproportionately affected population. |
| EU Agency for Fundamental Rights, April 2021, “Coronavirus Pandemic in the EU – Fundamental Rights Implications: Vaccine Rollout and Equality of Access In the EU” | Report | Europe | Only a third of EU Member States have prioritised detainees in their national vaccination strategies. In addition to age, vaccine deployment should also take into account specific vulnerabilities of certain population groups, including those who live in prisons. |
| Geana et al., April 2021, “COVID-19 vaccine hesitancy among women leaving jails: A qualitative study” | Empirical | Recently released women from Midwest USA | Interviews with 25 women who had left US prisons revealed that individuals who live and work in prisons often do not trust vaccines despite being at high risk of contracting COVID-19. This mistrust is fuelled by lack of knowledge and understanding of the disease and the vaccine, and suspicion of any authority involved in the development or administration of the vaccine. Health education delivered to this population should address mistrust, misinformation, and belief in conspiracy theories. |
| Hershow et al., April 2021, “Rapid Spread of SARS-CoV-2 in a State Prison After Introduction by Newly Transferred Incarcerated Persons — Wisconsin, August 14–October 22, 2020.” | Morbidity and Mortality Weekly Report | Wisconsin prison, US | This report documents the rapid spread of an outbreak of SARS-CoV-2 in one US prison. The authors argue that vaccination of prison staff and residents is important to control transmission and end outbreaks. They emphasise that vaccination of staff alone would have been unlikely to prevent the outbreak described, and call for the prioritisation of those living and working in prisons for vaccination. |
| Inside Time, July 2021, “Vaccine refusal rate over 50% at some jails” | News | England and Wales | Data from five prisons in England and Wales illustrate that between one- and two-thirds of people in prisons declined a first vaccine dose. Reasons include vaccine scepticism and belief in conspiracy theories. |
| Jacobsen et al., May 2021, “Care for Incarcerated Patients Hospitalized with COVID-19” | Narrative review | US prisons | Exclusion of people in prisons from COVID-19 vaccination trials prevents them from having early access to medication. Lack of prioritisation of this group in national vaccination plan has led some states to make their own decisions. |
| Jung and Han, July 2021, “The Effect of Knowledge, Attitudes, and Practices of Korean Correctional Officers about COVID-19 on Job Stress” | Empirical | South Korea | Health authorities in South Korea prioritised correctional facilities as part of their national vaccination programmes, especially for facilities where staff and residents work and live in close proximity. |
| Justice Lab, December 2020, “Recommendations for Prioritization and Distribution of COVID-19 Vaccine in Prisons and Jails” | Report | United States | This report recommends that states should prioritise vaccine distribution to all incarcerated people, to be delivered at the same time or earlier as vaccines for correctional officers (essential workers and first responders). It also suggests that medical and public health professionals should develop vaccine distribution and implementation plans that are specific to correctional systems, as well as ensure that state advisory groups include prison representatives, to ensure their priorities and needs are sufficiently met. |
| Khorasani et al., May 2021, “COVID-19 Vaccine Interest among Corrections Officers and People Who Are Incarcerated at Middlesex County Jail, Massachusetts” | Brief report | US prisons | This study found that the majority of people incarcerated in prisons, particularly those of Black ethnicity, were hesitant to receive a COVID-19 vaccine. Addressing misinformation and mistrust are key for overcoming hesitancy among this population. |
| Kowalski et al., 2020, “Jails in the Time of Coronavirus” | Empirical | US prisons | The authors advocate the following measures to address COVID-19 in prisons: education of staff and inmates, securing vaccination supplies, vaccinating early, using proactive steps, leadership from the public health sector to help with planning, and developing uniform directives for care. They also highlight the importance of building a relationship with the public, relying on credible resources, gathering supplies, having both a plan and sufficient training in place, and communicating with authorities. |
| Kronfli and Akiyama, December 2020, “Prioritizing incarcerated populations for COVID-19 vaccination and vaccine trials” | Commentary | US and Europe | People in correctional facilities should be considered high priority for COVID-19 vaccination because of their poor living conditions, a higher prevalence of chronic diseases compared to those in the community, and increased likelihood of transmitting the virus to staff and visitors. The paper also discusses failures to include incarcerated individuals in COVID-19 vaccine trials, citing historical medical exploitation, systemic barriers, ethical challenges, logistical reasons, and difficulties in ensuring representation of lived experience. It argues that because vaccination of incarcerated individuals is conditional upon successful vaccine trials, it is unethical to not offer trial participation to the incarcerated. Although incarcerated populations still need to be protected from coercion and exploitation, respect for the incarcerated also requires recognition of their autonomy in decision-making and respect for basic human rights. |
| Lambert and Wilkinson, January 2021, “Trust, efficacy and ethicacy when testing prisoners for COVID-19” | Discussion paper | UK, US and Europe | The authors highlight two key issues regarding vaccination of imprisoned people. First, there is a misconception that imprisoned people are young and therefore at low risk, as in reality many people in prisons are at risk because of underlying chronic conditions, age, and environment. Second, whilst many high-income countries were conducting national vaccination programmes for COVID-19, prison populations have not been considered in existing planning and guidance. In the UK, the Joint Committee on Vaccination and Immunisation’s stated priority for vaccines is to prevent deaths, protect health, and support social care staff and systems; it does not mention prisons. |
| Lewis et al., April 2021, “Community-Associated Outbreak of COVID-19 in a Correctional Facility — Utah, September 2020–January 2021” | Morbidity and Mortality Weekly Report. | US prisons | The authors report that of over 1,000 imprisoned people that tested positive for SARS-CoV-2 in a Utah prison, 31 were hospitalised and 12 died. Vaccination of imprisoned people would likely help prevent or limit the spread of infection in prisons. Nonetheless, preventing the introduction and spread of SARS-CoV-2 in prisons also depends on quarantine and implementation of available prevention measures. |
| Liebrenz et al., January 2021, “Prisoner's Dilemma: Ethical questions and mental health concerns about the COVID-19 vaccination and people living in detention” | Editorial | US prisons | This report concludes that vaccinating imprisoned people protects the whole community, and states that it is also important to recognise the high levels of mental illness seen in imprisoned people. This population may be particularly anxious about vaccination, and a lack of trust towards staff is probable. Healthcare staff must play a key role in educating imprisoned people and gaining their trust. The authors acknowledge that sometimes healthcare workers themselves are reluctant to be vaccinated. The question of how best to manage those who refuse to be vaccinated also needs to be considered, with a balance between public health efforts and individual rights. Global equity of vaccine distribution is another key issue. Many imprisoned people are from countries that do not have a robust public health system. |
| Macmadu and Brinkley-Rubinstein, January 2021, “Essential Strategies to Curb COVID-19 Transmission in Prisons and Jails” | Opinion | US | Disease outbreaks that occur in prisons rarely stay in prisons, and thus pose a risk to those who live and work in prisons but also to the wider community. The authors provide three key strategies to reduce the spread of COVID-19 in prisons: mass testing, prioritised vaccination, and decarceration.  Imprisoned people and staff working in prisons should be prioritised for vaccination. However, the authors report that specific US states have excluded imprisoned people from vaccination plans, citing high turnover and resulting inability to complete the two-vaccine schedule as a key reason for this. The authors argue that such a position is unacceptable, as those who are released prior to the second dose could be referred to community services for further vaccination. Partnership between prisons and local public health departments will be key to implement this policy. |
| Mills and Salisbury, December 2020, “The challenges of distributing COVID-19 vaccinations” | Commentary | United Kingdom | Written just prior to the availability of effective vaccines against COVID-19, the authors of this paper stress the need for transparency about vaccine distribution and implementation. The authors recognise those who live and work in prisons as a priority group, suggesting that in the UK, mobile teams should be employed for prison vaccination drives. They acknowledge the logistical problems in reaching 80,000 people in 117 prisons in England and Wales, and the need for careful planning to ensure that there is neither a shortage nor wastage of doses. |
| Neufeld et al., May 2021, “Prisons need to be included in global and national vaccinations effort against COVID-19” | Commentary | Worldwide | There is scarce data regarding which countries offer COVID-19 vaccination in prisons. Vaccine allocation is managed differently within and between countries. The authors cite a US example, where strategies differ between states, with some prioritising imprisoned people and others prioritising staff only. Considering the underlying health problems and environmental factors that increase risk of infection among those who live and work in prisons, failing to prioritise this group for vaccination violates the UN Standard Minimum Rules for the Treatment of Prisoners, and allows infection to spread between prisons and the community. |
| Novisky et al., April 2021, “Incarceration as a Fundamental Social Cause of Health Inequalities: Jails, Prisons and Vulnerability to COVID-19” | Article (Analysis/Debate) | US | The authors point out that although many experts across the world have called for imprisoned people to be prioritised for COVID-19 vaccination, there is limited data on the number of vaccines that have so far been distributed to this population. At the time of publication, neither the WHO nor most US state prison systems had released plans for vaccinating imprisoned people. National and local action should ensure that imprisoned people have timely access to vaccination. |
| Ramaswamy et al., June 2021, “Recommendations for Delivering COVID-19 Vaccine in Jails: Evidence from Kansas, Iowa, Nebraska, and Missouri” | Opinion | US | US jails have limited staff to implement a vaccination rollout. As such, collaboration between jail administrators, jail medical staff, and local health departments is required. Other issues that have prevented prioritisation of prison populations for vaccination include costs, short length of stay of imprisoned people, availability of medical staff to administer jabs, and safety concerns for staff. Vaccine information needs to be provided in a transparent, clear, and consistent manner to promote uptake. |
| Reiter, April 2021, “Does a Public Health Crisis Justify More Research with Incarcerated People?” | Essays | US | Despite ethical recommendations for the inclusion of incarcerated people in phase III vaccine trials, the author argues that the poor conditions in American prisons are too pervasive to permit either free and knowing consent to or non-coerced participation in vaccine trials. The author states that these structural risks, inherent to U.S. incarceration, seem likely to overwhelm any attempts to mitigate the individual risks of knowingly, freely, and safely participating in a randomised controlled vaccine or drug trial. |
| Ryckman et al., August 2021, “Outbreaks of COVID-19 variants in US prisons: a mathematical modelling analysis of vaccination and reopening policies” | Empirical | US | Even in prisons with low room occupancies, hospitalisation risks are substantial when prisons house medically vulnerable populations. Risks of large outbreaks (>20% of residents infected) are substantially higher if infections are repeatedly introduced. The paper suggests that after achieving high vaccine coverage, prisons with mostly one-to-two-person cells that have higher baseline immunity from previous outbreaks can resume in-person activities with lower risk of a widespread outbreak, provided they maintain widespread NPIs, continue testing, and take measures to protect the medically vulnerable. |
| Sanchez et al., 2020, “COVID-19 in prisons: an impossible challenge for public health?” | Opinion | Brazil | Influenza vaccination should be a key priority, as reducing the incidence of influenza will decrease the number of symptomatic people being tested for COVID-19. Information for health personnel and security staff, availability of PPE, diagnostic testing, influenza vaccination, and adjustment of COVID-19 risk prevention practices are indispensable. Preventive work leave for those belonging to the risk group should be granted. |
| Simas et al., March 2021, “For an equitable COVID-19 vaccination strategy for the population deprived of liberty” | Thematic section | Brazil and US | Political rhetoric disregards the fundamental rights of imprisoned populations to have access to the same level of care as the free population. Prioritisation of prison professionals for COVID-19 vaccination without prioritising the prison population is discriminatory, especially when they share the same environment. Although people in prisons were initially removed from the Brazilian national priority list for vaccination, they were subsequently included again due to a strong negative response from the public and a realisation of increased spread of the virus in prisons. |
| Simpson et al., April 2021, “Incarcerated people should be prioritised for covid-19 vaccination” | Editorial | Worldwide | The authors note that countries with the highest COVID-19 community transmission rates, including England, India, the US, and Australia, have not heeded the WHO’s advice to prioritise residents and staff in prisons for vaccination. This is due in part to conservative politicians and custodial staff representatives objecting to people who live in prisons being classified as a priority group. While several countries have prioritised older people with health conditions in prisons, the authors argue that this approach overlooks the unique risk factors that incarcerated people face, which place them and their contacts outside of prison at substantial risk of COVID-19 infection, transmission, and death. To address criticism, governments should communicate that vaccination in prisons benefits the wider community and that not doing so would violate human rights. |
| Stern et al., April 2021, “Willingness to Receive a COVID-19 Vaccination Among Incarcerated or Detained Persons in Correctional and Detention Facilities — Four States, September–December 2020” | Cross-sectional survey in 4 different USA states | USA | The authors conducted a cross-sectional survey of imprisoned people in four US states prior to a COVID-19 vaccination programme. They found that 45% of participants were willing to be vaccinated, although this was lower among participants who were younger and who identified as Black/African American. Among those who reported hesitancy to receive COVID-19 vaccination, many reported that they were awaiting more information or to see others vaccinated, as they had concerns about efficacy and/or safety. These individuals also expressed distrust of healthcare, correctional, or governmental personnel or institutions. Some thought vaccination was unnecessary as they did not feel they were at risk for COVID-19.  High rates of hesitancy highlight the need for culturally appropriate vaccination information, which needs to be delivered so that it can reach people of all levels of health literacy. This might include the use of peer influencers. Interventions to improve vaccine confidence should not be punitive. |
| Strassel et al., November 2020, “Covid-19 Vaccine Trials and Incarcerated People — The Ethics of Inclusion” | Perspective | US | The authors believe that researchers should take a more measured approach when considering whether to include incarcerated people in multisite vaccination trials. Correctional facilities have generally failed to meet minimum clinical care and public health standards. Benefits from taking part in trials are not guaranteed, and vaccines being trialled are more likely to fail than succeed. Furthermore, risks associated with research participation may be heightened in settings with limited or no on-site clinical resources for participants who have COVID-19 symptoms, complications, or severe adverse events.  The authors also argue that insufficient data exists regarding the spectrum of risks and benefits associated with current vaccine candidates, and that fast-paced COVID-19 trials might raise unexpected safety concerns. They also raise the issue of real or perceived pressure from authorities to enrol in research. Although some incarcerated people may be willing to take on substantial risk to receive an experimental vaccine, desperation or fear of COVID-19 may lead others to underestimate the level of risk involved. |
| Strodel et al., June 2021, “COVID-19 vaccine prioritization of incarcerated people relative to other vulnerable groups: An analysis of state plans” | Research article | US | The authors found that incarcerated people were consistently not prioritised in the first phase of the vaccine rollout, while other vulnerable groups who shared similar environmental risks were prioritised. They argue that this discrepancy highlights a potential violation of human rights regarding equitable access to care for incarcerated people. While early prioritisation of correctional staff may protect people who are incarcerated by reducing community-to-facility transmission, authors expressed concern that these workers are prioritised earlier for immunisation despite being exposed to less daily environmental risks than incarcerated people.  Analysis shows that incarcerated people, regardless of age, are often given lower priority than individuals 65 years and older in the community. This presents an ethical concern, as incarcerated older individuals are a particularly vulnerable population who often experience delays in routine and specialty medical care, including treatment for COVID-19. Older adults in the community are able to receive care sooner, via emergency medical service providers.  The review also identified that although states’ COVID-19 vaccination strategies often emphasised correctional leaders as key partners, this partnership was poorly defined. The authors call for further research to explore the nature of partnerships between public health and correctional leaders in the rollout of COVID-19 vaccines. Lack of support for the logistical components of vaccine rollout in carceral settings could lead to further de-prioritisation of incarcerated people, as convenience could influence allocation decisions. Aside from a lack of clear guidance on vaccination rollout by correctional staff, vaccine hesitancy among this group may also hinder efforts to curb COVID-19 in carceral settings. |
| Tavoschi et al., March 2021, “Equitable and tailored access to covid-19 vaccine for people in prison” | Opinion | Europe | The authors propose that prison staff are essential workers, and that absences should be minimised to enable prisons to function safely and humanely. Vaccination is a key strategy to minimise absenteeism. Prison populations should also be prioritised for vaccination due to their high burden of underlying disease, including non-communicable diseases known to correlate with low socioeconomic status. Those in prison embody the COVID syndemic concept, i.e., that biological, economic, and social factors exacerbate the interaction between non-communicable diseases and COVID-19 to increase susceptibility to infection and poor health outcomes.  Implementing SARS-CoV-2 vaccination programmes in prisons presents additional challenges, which include suboptimal healthcare delivery and information systems, and poor links to community systems. Evidence on how to design and implement prison vaccination services is scarce, as operational research on prison health is usually not high on national and international public health agendas. |
| UK Joint Committee on Vaccination and Immunisation, March 2021, “Letter from the JCVI to the Health and Social Care Secretary on further considerations on phase 1 advice: 1 March 2021” | Letter | UK | The committee acknowledges that prison officers may be sources of transmission in a COVID-19 outbreak in prison settings, and that officers work with individuals vulnerable to COVID-19, yet they do not consider officers to have the same level of risk of exposure as health and social care workers. The committee found it difficult to recommend additional prioritisation of detainees based only on potential for increased risk of exposure in a detained setting. |
| SAGE EMG Transmission Group, March 2021, “COVID-19 Transmission in Prison Settings” | Report | UK | Prisons have limited primary healthcare teams to deliver testing, vaccination, and other infection control interventions in outbreaks, in addition to delivering primary healthcare services. Teams are often depleted due to staff cases of COVID-19 or isolating contacts. This impacts the operational capacity to manage outbreaks.  Restricting vaccination to all prison residents and staff over the age of 50 was found to be considerably less effective at preventing outbreaks. Reverse cohorting may continue to be necessary. The authors recommend routine staff testing, particularly for unvaccinated staff, to prevent the virus being imported into prisons. |
| Wang et al., September 2020, Ethical Considerations for COVID-19 Vaccine Trials in Correctional Facilities. | Viewpoint | US | Prisons and jails are exempted from participating in COVID-19 vaccine trials. This omission, according to the authors, is an example of the unintended consequences of well-intentioned policies. The authors propose the following: obtaining input from currently and formerly incarcerated individuals and from those who work in prisons; recruiting from minority groups to improve the external validity of COVID-19 vaccine trials, which tend to recruit mostly White participants; ensuring receipt of efficacious vaccines and care after trials conclude; and deeper reflection on the implementation of vaccines in correctional systems. |
| WHO, February 2021, “Preparedness, prevention and control of COVID-19 in prisons and other places of detention” | Interim guidance | Europe | This guidance argues that, in line with the Mandela Rules, prison administrations should take into account =the individual needs of people in prison, especially the most vulnerable. It calls for measures to protect and promote the rights of those prison with special needs. It reiterates that healthcare provision in prisons is the responsibility of the state and should meet the same standards that are available in the community, without any charge. Finally, it calls for prioritisation of vaccines for healthcare professionals, non-healthcare staff that provide services carrying significant risk of infection, older adults, and individuals at higher risk of death due to underlying conditions such as heart disease and diabetes. |
| WHO, 2021, “Why people living and working in detention facilities should be included in national COVID-19 vaccination plans” | Advocacy brief | Europe | Based on various case studies from European countries, this brief highlights how the limited resources available for the prison population, including access to testing and personal protective equipment (PPE), present challenges for curbing the spread of COVID-19 in prisons. Vaccination of the prison population and workforce should be a key public health measure. The brief advocates that prison staff and prison healthcare staff are essential health and care workers and should be prioritised for vaccines. It also suggests that the cut-off age for the elderly should be lowered to 50 years because of the poor health status of people in detention and the ageing effect of prison itself.  The brief proposes a robust information system that is capable of recording and tracking those receiving their first dose in the community and then needing their second in detention, and vice versa. Access to PPE and to rapid diagnostic tests should be ensured in all areas of the criminal justice system. Good governance and a coordinated response between public health services and the ministries responsible for health in prisons are imperative. |
