## Supplementary material for "COVID-19 vaccine for people who live and work in prisons worldwide: A scoping review": Quality assessments of the included studies

**Appendix 3 – Quality assessments of the included studies**

Table 1 – Observation, cohort, and cross-sectional studies

|  |  |  | Studies |  |  |  |  |
| --- | --- | --- | --- | --- | --- | --- | --- |
| Checklist | **Chin et al., 2021b** | **Hershow et al., 2020** | **Jung and**  **Han, 2021** | **Khorasani et al. (2021)** | **Ramaswamy et al. (2021)** | **Lewis et al., 2021** | **Stern et al., 2021** |
| 1. Was the research question or objective in this paper clearly stated? | Yes | Yes | Yes | Yes | Yes | Yes | Yes |
| 2. Was the study population clearly specified and defined? | Yes | Yes | Yes | Yes | Yes | Yes | Yes |
| 3. Was the participation rate of eligible persons at least 50%? | N/A | No | Yes | Yes | Yes | No | No |
| 4. Were all the subjects selected or recruited from the same or similar populations (including the same time period)? Were inclusion and exclusion criteria for being in the study prespecified and applied uniformly to all participants? | N/A | Yes | Yes | Yes | Yes | Yes | Yes |
| 5. Was a sample size justification, power description, or variance and effect estimates provided? | N/A | No | Yes | N/A | N/A | No | No |
| 6. For the analyses in this paper, were the exposure(s) of interest measured prior to the outcome(s) being measured? | Yes | Yes | Yes | N/A | N/A | Yes | N/A |
| 7. Was the timeframe sufficient so that one could reasonably expect to see an association between exposure and outcome if it existed? | Yes | Yes | Yes | N/A | N/A | Yes | N/A |
| 8. For exposures that can vary in amount or level, did the study examine different levels of the exposure as related to the outcome (e.g., categories of exposure, or exposure measured as continuous variable)? | N/A | N/A | Yes | N/A | N/A | N/A | N/A |
| 9. Were the exposure measures (independent variables) clearly defined, valid, reliable, and implemented consistently across all study participants? | Yes | Yes | No | Yes | Yes | Yes | Yes |
| 10. Was the exposure(s) assessed more than once over time? | N/A | Yes | No | No | No | Yes | No |
| 11. Were the outcome measures (dependent variables) clearly defined, valid, reliable, and implemented consistently across all study participants? | Yes | Yes | No | N/A | N/A | Yes | Yes |
| 12. Were the outcome assessors blinded to the exposure status of participants? | No | No | N/A | N/A | No | No | No |
| 13. Was loss to follow-up after baseline 20% or less? | N/A | N/A | Yes | N/A | N/A | N/A | N/A |
| 14. Were key potential confounding variables measured and adjusted statistically for their impact on the relationship between exposure(s) and outcome(s)? | Yes | N/A | No | Yes | Yes | N/A | Yes |
| Quality Rating | Cannot decide | Fair | Poor | Fair | Good | Fair | fair |

Table 2 – Qualitative papers

| Checklist | | Geana et al., 2021 |
| --- | --- | --- |
| Section A: Are the results valid? | 1. Was there a clear statement of the aims of the research? | Yes |
|  | 2. Is a qualitative methodology appropriate? | Yes |
|  | 3. Was the research design appropriate to address the aims of the research? | Yes |
|  | 4. Was the recruitment strategy appropriate to the aims of the research? | Cannot tell |
|  | 5. Was the data collected in a way that addressed the research issue? | Yes |
|  | 6. Has the relationship between researcher and participants been adequately considered? | Cannot tell |
| Section B: What are the results? | 7. Have ethical issues been taken into consideration? | Yes |
|  | 8. Was the data analysis sufficiently rigorous? | Yes |
|  | 9. Is there a clear statement of findings? | Yes |
| Section C: Will the results help locally? | 10. How valuable is the research? | Yes |
|  | Quality Rating | Good |

Table 3 – Secondary data analysis

| Checklist | Kowalski et al., 2020 | Strassle et al., 2020 | Strodel et al., 2021 | Wang et al., 2020 |
| --- | --- | --- | --- | --- |
| Does the paper address a clear issue? | Yes | Yes | Yes | Yes |
| Are the recommendations valid? | Yes | Yes | Yes | Yes |
| Does the paper provide details on its methodology? | Yes | N/A | Yes | No |
| Is the paper likely to account for important recent developments, i.e., how up to date is it? | Yes | Yes | Yes | Yes |
| Has the paper been subject to peer review and testing? | Yes | Yes | Yes | Yes |
| Is there a conflict of interest in the development and publication of these guidelines? | No | No | Yes | No |
| Are practical recommendations made? | Yes | Yes | Yes | Yes |
| Are the recommendations strong? | Yes | Yes | Yes | Yes |
| Is the research valuable? | Yes | Yes | Yes | Yes |
| Quality Rating | Good | Good | Good | Good |

Table 4 – Cost-effectiveness analyses

| Checklist | Blumberg et al., 2021 | Chapman et al., 2021 | Ryckman et al., 2021 |
| --- | --- | --- | --- |
| Identify the study as an economic evaluation or use more specific terms such as “cost-effectiveness analysis”, and describe the interventions compared. | No | No | No |
| Provide a structured summary of objectives, perspective, setting, methods (including study design and inputs), results (including base case and uncertainty analyses), and conclusions. | No | Yes | Yes |
| Provide an explicit statement of the broader context for the study. | Yes | Yes | Yes |
| Present the study question and its relevance for health policy or practice decisions. | Yes | Yes | Yes |
| Describe characteristics of the base case population and subgroups analysed, including why they were chosen. | Yes | Yes | Yes |
| State relevant aspects of the system(s) in which the decision(s) need(s) to be made. | Yes | Yes | Yes |
| Describe the perspective of the study and relate this to the costs being evaluated. | N/A | N/A | N/A |
| Describe the interventions or strategies being compared and state why they were chosen. | Yes | Yes | Yes |
| State the time horizon(s) over which costs and consequences are being evaluated and say why appropriate. | No | Yes | Yes |
| Report the choice of discount rate(s) used for costs and outcomes and say why appropriate. | N/A | N/A | N/A |
| Describe what outcomes were used as the measure(s) of benefit in the evaluation and their relevance for the type of analysis performed. | Yes | Yes | Yes |
| Single study-based estimates: Describe fully the design features of the single effectiveness study and why the single study was a sufficient source of clinical effectiveness data. | No | Yes | Yes |
| Synthesis-based estimates: Describe fully the methods used for identification of included studies and synthesis of clinical effectiveness data. | N/A | N/A | N/A |
| If applicable, describe the population and methods used to elicit preferences for outcomes. | N/A | N/A | N/A |
| Single study-based economic evaluation: Describe approaches used to estimate resource use associated with the alternative interventions. Describe primary or secondary research methods for valuing each resource item in terms of its unit cost. Describe any adjustments made to approximate to opportunity costs. | N/A | N/A | N/A |
| Model-based economic evaluation: Describe approaches and data sources used to estimate resource use associated with model health states. Describe primary or secondary research methods for valuing each resource item in terms of its unit cost. Describe any adjustments made to approximate to opportunity costs. | N/A | N/A | N/A |
| Report the dates of the estimated resource quantities and unit costs. Describe methods for adjusting estimated unit costs to the year of reported costs if necessary. Describe methods for converting costs into a common currency base and the exchange rate. | N/A | N/A | N/A |
| Describe and give reasons for the specific type of decision analytical model used. Providing a figure to show model structure is strongly recommended. | Yes | Yes | Yes |
| Describe all structural or other assumptions underpinning the decision-analytical model. | Yes | Yes | Yes |
| Describe all analytical methods supporting the evaluation. This could include methods for dealing with skewed, missing, or censored data; extrapolation methods; methods for pooling data; approaches to validate or make adjustments (such as half cycle corrections) to a model; and methods for handling population heterogeneity and uncertainty. | Yes | Yes | Yes |
| Report the values, ranges, references, and, if used, probability distributions for all parameters. Report reasons or sources for distributions used to represent uncertainty where appropriate. Providing a table to show the input values is strongly recommended. | Yes | Yes | Yes |
| For each intervention, report mean values for the main categories of estimated costs and outcomes of interest, as well as mean differences between the comparator groups. If applicable, report incremental cost-effectiveness ratios. | Yes | Yes | Yes |
| Single study-based economic evaluation: Describe the effects of sampling uncertainty for the estimated incremental cost and incremental effectiveness parameters, together with the impact of methodological assumptions (such as discount rate, study perspective). | N/A | N/A | N/A |
| Model-based economic evaluation: Describe the effects on the results of uncertainty for all input parameters, and uncertainty related to the structure of the model and assumptions. | Yes | Yes | Yes |
| If applicable, report differences in costs, outcomes, or cost-effectiveness that can be explained by variations between subgroups of patients with different baseline characteristics or other observed variability in effects that are not reducible by more information. | No | Yes | Yes |
| Summarise key study findings and describe how they support the conclusions reached. Discuss limitations and the generalisability of the findings and how the findings fit with current knowledge. | Yes | Yes | Yes |
| Describe how the study was funded and the role of the funder in the identification, design, conduct, and reporting of the analysis. Describe other non-monetary sources of support. | Yes | Yes | Yes |
| Describe any potential for conflict of interest of study contributors in accordance with journal policy. In the absence of a journal policy, we recommend authors comply with International Committee of Medical Journal Editors recommendations. | Yes | Yes | Yes |
